# Evaluation of a SARS-CoV-2 surrogate virus neutralization test for detection of antibody in human, canine, cat and hamster sera

**DOI:** 10.1101/2020.07.28.20163592

**Authors:** Ranawaka APM Perera, Ronald Ko, Owen TY Tsang, Christopher J Brackman, Esther M. W. To, Hui-ling Yen, Kathy Leung, Samuel MS Cheng, Kin Ho Chan, Karl CK Chan, Ka-Chi Li, Linda Saif, Vanessa R. Barrs, Joseph T Wu, Thomas H. C. Sit, Leo LM Poon, Malik Peiris

## Abstract

Surrogate neutralization assays for SARS-CoV-2 that can be done without biosafety-level-3 containment and across multiple species are desirable. We evaluate a recently developed surrogate virus neutralization test (sVNT) in comparison to 90% plaque reduction neutralization tests (PRNT90) in human, canine, cat and hamster sera and found excellent concordance between the two assays. Using a panel of immune sera to other coronaviruses, we confirm the lack of cross reactivity in sVNT and PRNT90 assays.

## Introduction

SARS-CoV-2 virus emerged in 2019 to cause a pandemic. It is important to detect antibody responses in humans and animals to ascertain evidence of past infection with SARS-CoV-2. Antibody assays that are transferable across species are desirable because SARS-CoV-2 infects pets and other farmed animals (e.g. mink) [1,2,3] and for monitoring antibody responses in experimental animal models [4]. ELISA assays to the virus spike receptor binding domain (RBD) which has least cross-reactive epitopes with other coronaviruses, the whole spike protein or nucleoprotein are widely used for detection of antibody in humans [5,6,7]. Virus neutralization assays are used for confirming positive results but require handling live virus in biosafety level 3 (BSL-3) containment or a pseudotyped virus.

ELISA testing for antibody in other animal species requires assays to be re-optimized with the relevant species-specific anti-Ig-conjugate for detecting immunoglobulins of each species. For some species relevant anti-Ig reagents may not be available. The currently available generic alternative has been to use virus neutralization tests which usually involves handling live virus in BSL-3 containment. A “surrogate” virus neutralization test (sVNT) that can be done in BSL-2 containment has recently been reported [8]. It is an assay that relies on competitive inhibition of the interaction of ACE-2 receptor coated on an ELISA plate with horse radish peroxidase labelled virus spike receptor binding domain. We used a panel of sera from patients or animals with RT-PCR confirmed SARS-CoV-2 infection and corresponding controls to evaluate this sVNT assay in comparison to the “gold-standard” PRNT90 test.

## Materials and Methods

### Sera

Sera (n=43) from SARS-CoV-2 RT-PCR confirmed patients and negative controls (n-42) collected from blood donors in 2017 were used [6]. Dogs (n=4) and a cat with RT-PCR confirmed SARS-CoV-2 were included in the study (4). Control dog sera were obtained from a study of canine influenza in 2017 [9] and sera from cats submitted for routine veterinary diagnostic testing in 2020 were used as controls. Hamster sera from experimentally infected hamsters and controls were from a previous study [4].

### Coronavirus immune sera

Immune sera specific for alphacoronaviruses (porcine respiratory coronavirus, feline infectious peritonitis virus, canine coronavirus and porcine transmissible gastroenteritis virus), betacoronaviruses (SARS-coronavirus) and gamma-coronavirus (infectious bronchitis virus) were obtained from BEI Resources (animal CoV reagents supplied to BEI by Dr Linda Saif (http://www.beiresources.org/About/BEIResources.aspx) or generated by Dr Linda Saif (Table 2). Dr Stanley Perlman provided mouse serum to mouse hepatitis virus (strains A59 and JHM) (Table 1). The homologous antibody titres to the immunising virus were also provided by Dr Linda Saif or Dr Stanley Perlman (Table 2).

**Table 1:** Performance characteristics of the surrogate virus neutralization (sVN) test in comparison with 90% plaque reduction neutralization test.

|  | **sVNT pos / PRNT90 pos**<br>**PRNT titers and sVN % inhibition** | **sVNT neg /**<br>**PRNT90 neg** | **sVNT / PRNT90**<br>**discrepant** | **Total** |
| --- | --- | --- | --- | --- |
| **SARS-CoV-2 infected dogs** | 3<br>(PRNT titres 40, 80, 160<br>sVNT %inhibition 33%, 66%, 78%) | 1 (acute serum,<br>later<br>seroconverted) | 0 | 4 |
| **Control dogs** | 0 | 40 | 0 | 40 |
| **SARS-CoV-2 infected cats** | 1 (PRNT titer320; sVNT % inhibition 83%) | 0 | 0 | 1 |
| **Control cats** | 0 | 19 | 0 | 19 |
| **SARS-CoV-2 infected**<br>**hamsters** | 3<br>(PRNT titers 320, 640, 640; sVNT % inhibition<br>83%; 83%; 83%) | 0 | 0 | 3 |
| **Control hamsters** | 0 | 2 | 0 | 2 |
| **SARS-CoV-2 infected human sera** |  |  |  |  |
| **<10 days after illness onset** | 3# | 3 | 0 | 6 |
| **≥10 days after illness onset** | 37# | 0 | 0 | 37 |
| **Sera from humans collected**<br>**in 2017** | 0 | 42 | 0 | 42 |
| **Total** | **47** | **107** | **0** | **154** |

**Table 2:**
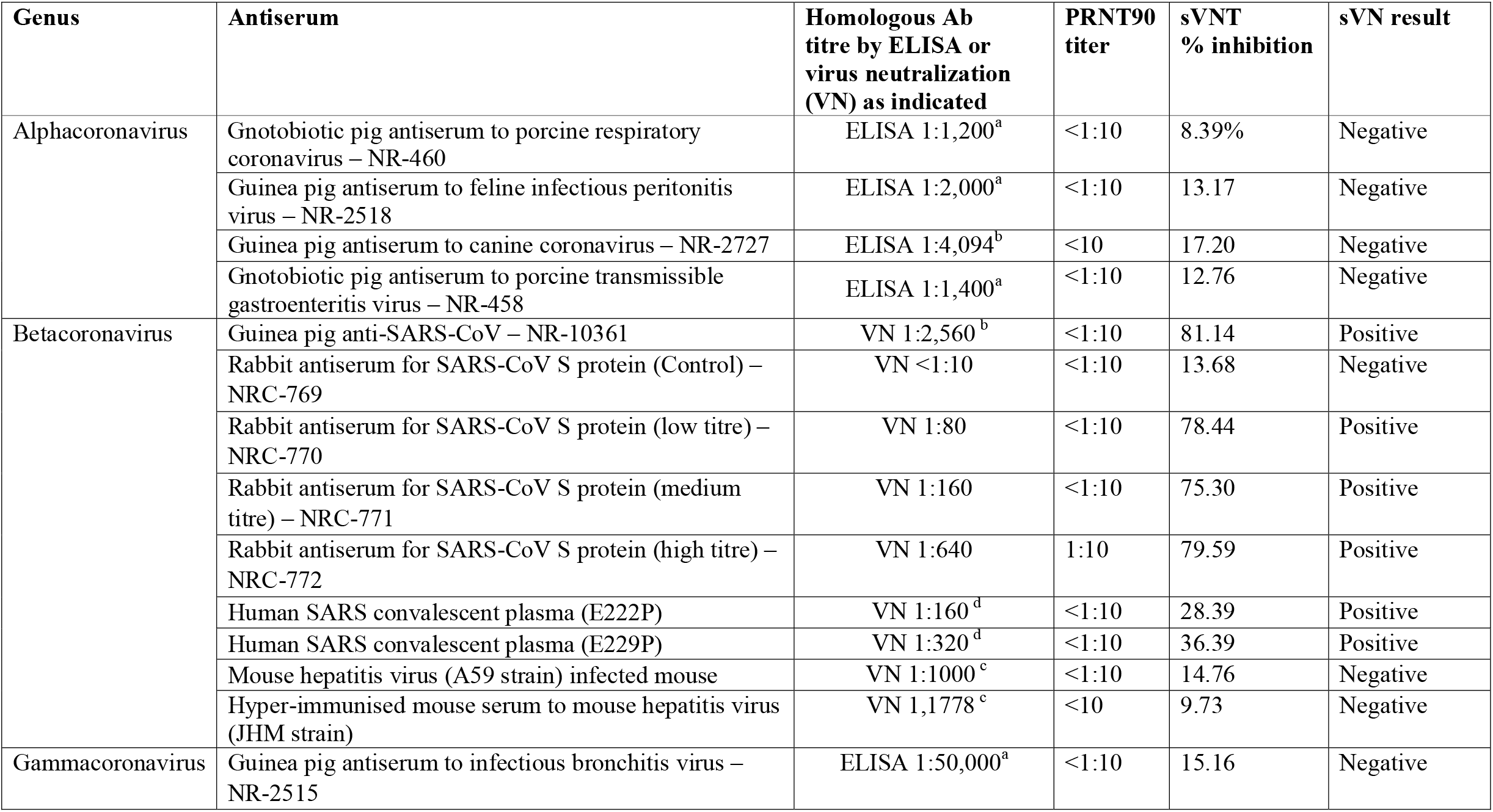
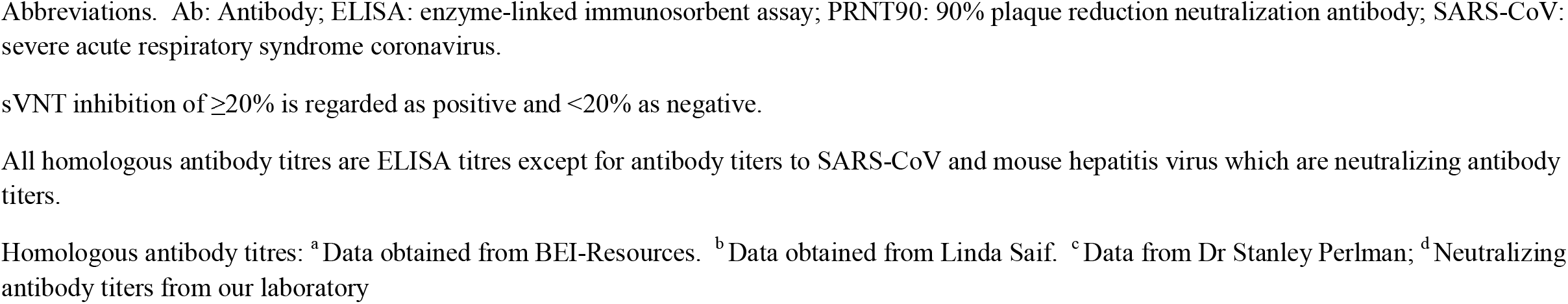
SARS-CoV-2-CoV surrogate virus neutralization (sVN) and 90% plaque neutralization (PRNT90) cross-reactivity with immune sera to other coronaviruses.

| Genus | Antiserum | Homologous Ab titre by ELISA or virus neutralization (VN) as indicated | PRNT90 titer | sVNT % inhibition | sVN result |
| --- | --- | --- | --- | --- | --- |
| Alphacoronavirus | Gnotobiotic pig antiserum to porcine respiratory coronavirus – NR-460 | ELISA 1:1,200<sup>a</sup> | <1:10 | 8.39% | Negative |
|  | Guinea pig antiserum to feline infectious peritonitis virus – NR-2518 | ELISA 1:2,000<sup>a</sup> | <1:10 | 13.17 | Negative |
|  | Guinea pig antiserum to canine coronavirus – NR-2727 | ELISA 1:4,094<sup>b</sup> | <10 | 17.20 | Negative |
|  | Gnotobiotic pig antiserum to porcine transmissible gastroenteritis virus – NR-458 | ELISA 1:1,400<sup>a</sup> | <1:10 | 12.76 | Negative |
| Betacoronavirus | Guinea pig anti-SARS-CoV – NR-10361 | VN 1:2,560<sup>b</sup> | <1:10 | 81.14 | Positive |
|  | Rabbit antiserum for SARS-CoV S protein (Control) – NRC-769 | VN <1:10 | <1:10 | 13.68 | Negative |
|  | Rabbit antiserum for SARS-CoV S protein (low titre) – NRC-770 | VN 1:80 | <1:10 | 78.44 | Positive |
|  | Rabbit antiserum for SARS-CoV S protein (medium titre) – NRC-771 | VN 1:160 | <1:10 | 75.30 | Positive |
|  | Rabbit antiserum for SARS-CoV S protein (high titre) – NRC-772 | VN 1:640 | 1:10 | 79.59 | Positive |
|  | Human SARS convalescent plasma (E222P) | VN 1:160<sup>d</sup> | <1:10 | 28.39 | Positive |
|  | Human SARS convalescent plasma (E229P) | VN 1:320<sup>d</sup> | <1:10 | 36.39 | Positive |
|  | Mouse hepatitis virus (A59 strain) infected mouse | VN 1:1000<sup>c</sup> | <1:10 | 14.76 | Negative |
|  | Hyper-immunised mouse serum to mouse hepatitis virus (JHM strain) | VN 1,1778<sup>c</sup> | <10 | 9.73 | Negative |
| Gammacoronavirus | Guinea pig antiserum to infectious bronchitis virus – NR-2515 | ELISA 1:50,000<sup>a</sup> | <1:10 | 15.16 | Negative |

Abbreviations. Ab: Antibody; ELISA: enzyme-linked immunosorbent assay; PRNT90: 90% plaque reduction neutralization antibody; SARS-CoV: severe acute respiratory syndrome coronavirus.
sVNT inhibition of $\geq 20\%$ is regarded as positive and $< 20\%$ as negative.
All homologous antibody titres are ELISA titres except for antibody titers to SARS-CoV and mouse hepatitis virus which are neutralizing antibody titers.
Homologous antibody titres: <sup>a</sup>Data obtained from BEI-Resources. <sup>b</sup>Data obtained from Linda Saif. <sup>c</sup>Data from Dr Stanley Perlman; <sup>d</sup>Neutralizing antibody titers from our laboratory

### Serological tests

The SARS-CoV-2 surrogate virus neutralization test kits were obtained from GeneScript USA, Inc, New Jersey, and the test carried out according to the manufacturer’s instructions. The test sera (60uL), positive or negative controls were diluted at 1:10 and mixed with an equal volume of horse radish peroxidase (HRP) conjugated to SARS-CoV-2 spike receptor binding domain (RBD) and incubated for 30 minutes at 37°C. Then, 100 μL each mix was added to the wells on the microtiter plate coated with ACE-2 receptor, the plate was sealed and incubated at 37°C for 15 minutes. The plates were then washed with wash-solution, tapped dry and 100 μL of 3,3’,5,5’-Tetramethylbenzidine (TMB) solution was added to each well and incubated in the dark at room temperature for 15 minutes. The reaction was stopped by addition of 50μL of Stop Solution to each well and the absorbance read at 450 nm in an ELISA reader. Assuming the positive and negative controls gave the recommended OD_450_ values, the % inhibition of each serum was calculated as (1-OD value of the sample / OD value of the negative control) x100. An inhibition of ≥20% is regarded as a positive result while that <20% is negative [8].

SARS-CoV-2 90% plaque reduction neutralization titer was determined as previously described [6].

## Ethical statement: Ethical statement

Collection of serum from patients with COVID-19 and healthy blood donor sera in 2017 was approved by the Research Ethics Committee of the Kowloon West Cluster reference No. KW/EX-20-039 (144-27) of the Hospital Authority of Hong Kong and the Institutional Review Board of The Hong Kong University and the Hong Kong Island West Cluster of Hospitals (IRB reference number UW16-254), respectively. Canine sera were residual sera from sera collected as part of routine clinical care and the Committee on the Use of Live Animals in Teaching and Research (CULATR) has waived animal ethics approval. Cat sera were residual sera collected routine diagnosis. The experimental study on hamsters was approved by the Committee on the Use of Live Animals in Teaching and Research, The University of Hong Kong (CULATR # 5323-20).

## Results and Discussion

Human (n=85), dog (n=44), cat (n=20) and hamster (n=5) sera from SARS-CoV-2 infected or controls, with known plaque reduction neutralization titres results, were tested in the sVNT assay. The concordance between the sVNT test and PRNT90 test was 100%, with all 47 PRNT90 positive sera also being positive in the sVNT assay while all 107 PRNT90 negative sera were negative in the sVNT (Table 1). Three of 4 sera collected from two RT-PCR positive dogs were positive in both sVNT and PRNT90 assays, the negative serum being an early serum collected from a dog which subsequently seroconverted by both assays. All 37 human sera from RT-PCR confirmed human patients obtained 10 or more days after onset of illness were positive in both assays whereas 3 of 6 sera collected prior to day 10 after illness onset were sero-negative in both assays. The sVNT assay is not meant to be quantitative. However, there was a semi-quantitative relationship between the % inhibition in the sVNT assay and PRNT90 titres (Figure).

**Figure:**
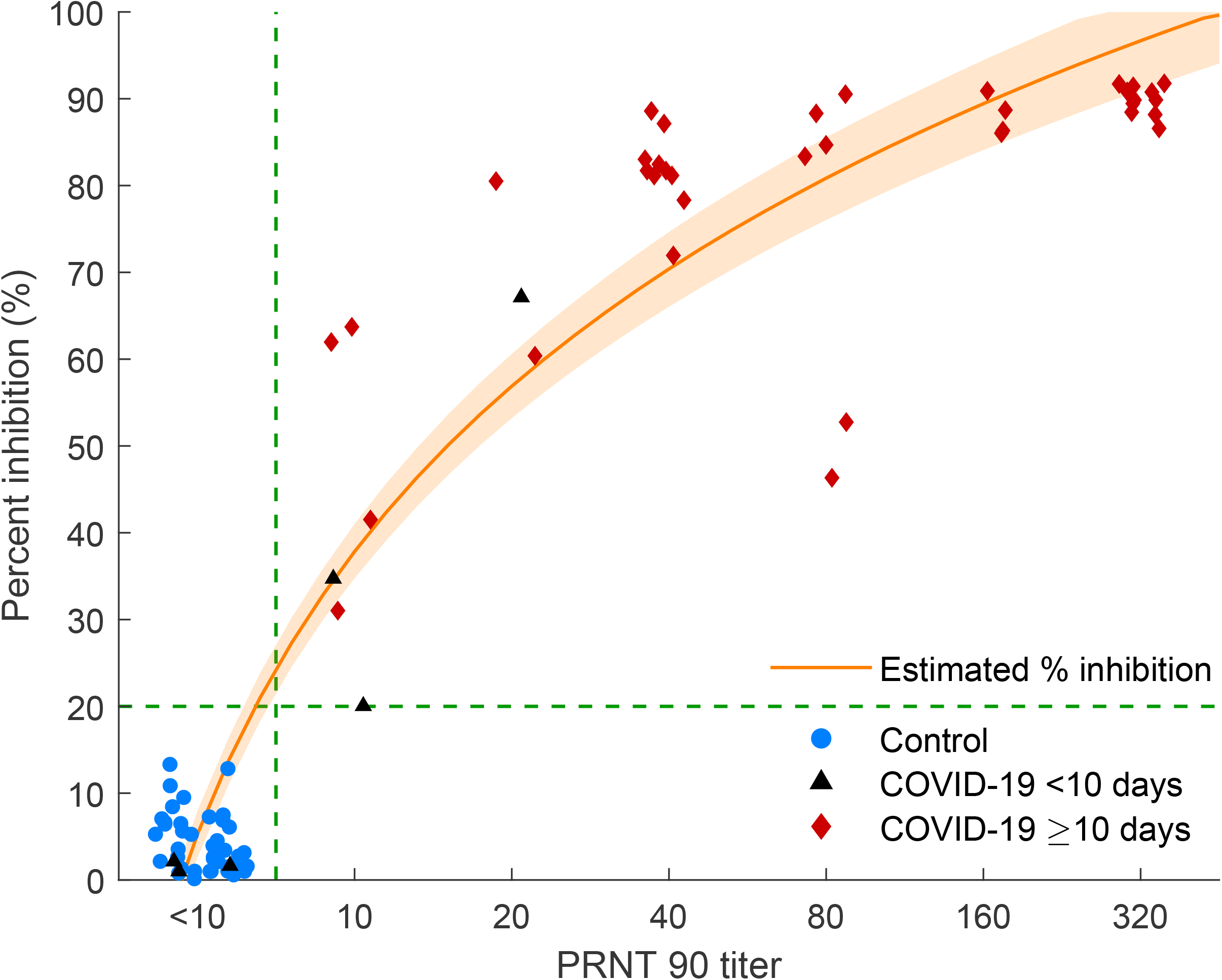
Correlation between 90% plaque reduction neutralization test (PRNT 90) titer and the percent inhibition in the surrogate virus neutralization test (sVNT). We fit a linear-log regression model between the percent inhibition in the sVNT assay and log-transformed PRNT90 titration titers. The orange line showed the estimated percent of inhibition and the orange shades showed the 95% confidence interval. Blue dots showed the control samples. Black triangles showed samples from confirmed COVID-19 cases obtained less than 10 days since symptom onset and red diamonds showed samples from confirmed cases obtained at least 10 days after symptom onset.

We investigated cross reactivity of immune sera to a range of alpha, beta and gamma coronaviruses in the sVNT and PRNT90 assays including antisera to feline infectious peritonitis virus, canine coronavirus, mouse hepatitis virus and SARS-CoV (Table 2). Two human SARS-CoV convalescent plasma samples were also included in this assessment. Cross reactivity in the sVNT assay was only detected with SARS convalescent human plasma (homologous PRNT90 titers of 1:160 and 1:320) and with a Guinea pig immune sera to SARS-CoV with a homologous neutralization titer of 1:2,560. Cross reactivity in the SARS-CoV-2 PRNT90 test was only observed with the high titre hyperimmune rabbit sera to SARS-CoV (homologous antibody titer 1:640).

We found excellent concordance between the sVNT and the “gold-standard” PRNT90 assays for SARS-CoV-2 antibody detection in humans, dogs, cat and hamster sera. It is likely that the sVNT assay can be used to detect SARS-CoV-2 antibody in any species in a highly specific and convenient manner. This assay would be of great utility as a species independent assay for primary testing for SARS-CoV antibodies in humans of animals or for independent confirmation of sera giving positive ELISA antibody results. While there was an overall correlation between PRNT90 titers and % inhibition in the sVNT assay, antibody quantitation of sVNT remains a challenge. We did not compare its performance with pseudo-particle neutralization assays for antibody. The sVNT test has the advantage of technical simplicity, speed, and not requiring cell culture facilities and BSL-3 containment.

## Data Availability

Data is stated in the manuscript

## Acknowledgments

We acknowledge research funding from the Health and Medical Research Fund, the Food and Health Bureau, The Government of Hong Kong Special Administration Region. We thank Dr Stanley Perlmann for the mouse hepatitis virus immune sera and relevant serological data. We thank Linfa Wang at Duke-NUS and GenScript for advice on the sVNT.

